# Prevalence and outcome of Covid-19 infection in cancer patients: a national VA study

**DOI:** 10.1101/2020.08.21.20177923

**Authors:** Nathanael R. Fillmore, Jennifer La, Raphael E. Szalat, David P. Tuck, Vinh Nguyen, Cenk Yildirim, Nhan V. Do, Mary T. Brophy, Nikhil C. Munshi

## Abstract

**Background:** Emerging data suggest variability in susceptibility and outcome to Covid-19 infection. Identifying the risk-factors associated with infection and outcomes in cancer patients is necessary to develop healthcare recommendations.

**Methods:** We analyzed electronic health records of the US National Veterans Administration healthcare system and assessed the prevalence of Covid-19 infection in cancer patients. We evaluated the proportion of cancer patients tested for Covid-19 and their confirmed positivity, with clinical characteristics, and outcome, and stratified by demographics, comorbidities, cancer treatment and cancer type.

**Results:** Of 22914 cancer patients tested for Covid-19, 1794 (7.8%) were positive. The prevalence of Covid-19 was similar across all ages. Higher prevalence was observed in African-American (AA) (15%) compared to white (5.5%; P<.001), in Hispanic vs non-Hispanic population and in patients with hematologic malignancy compared to those with solid tumors (10.9% vs 7.7%; P<.001). Conversely, prevalence was lower in current smoker patients, patients with other co-morbidities and having recently received cancer therapy (< 6 months). The Covid-19 attributable mortality was 10.9%. Highest mortality rates were observed in older patients, those with renal dysfunction, higher Charlson co-morbidity score and with certain cancer types. Recent (< 6 months) or past treatment did not influence mortality. Importantly, AA patients had 3.5-fold higher Covid-19 attributable hospitalization, however had similar mortality rate as white patients.

**Conclusion:** Pre-existence of cancer affects both susceptibility to Covid-19 infection and eventual outcome. The overall Covid-19 attributable mortality in cancer patients is affected by age, co-morbidity and specific cancer types, however, race or recent treatment including immunotherapy does not impact outcome.

**Fundings:** VA Office of Research and Development and National Institutes of Health.

## Introduction

The Covid-19 infection first reported in China, in December 2019^1^, has now spread worldwide affecting all demographics and regions. The emerging data suggests variability in susceptibility to the infection and ultimately outcome. A number of patient related factors, socio economic conditions, racial and ethnic differences and several comorbidities including obesity, diabetes and cardiovascular diseases have been associated with higher susceptibility and/or risk of mortality.^2-5^ The relatively higher transmission rate and associated greater risk of adverse outcome has highlighted the need to understand the epidemiologic characteristics of Covid-19 prevalence, and the risk factors associated with poor outcome and death, in order to establish the best possible public health policies. Cancer patients are considered to be at a higher risk of infections. This risk varies with functional status of the patient, the cancer type, and/or treatment modalities utilized.^6,7^ Thus, along with reducing exposure to the virus, other prophylactic as well as cancer-related risk factors may need to be addressed to decrease susceptibility to Covid-19 infection or to mitigate related complications in cancer patients. Small epidemiologic studies mainly from China and USA have also reported increased rates of death in cancer patients related to Covid-19.^8,9^ These observations have informed some changes and re-organization of cancer care worldwide^10^ but larger studies are needed to understand the comprehensive cancer-related issues with Covid-19 infection. Here we investigated the prevalence and outcome of Covid-19 infection among cancer patients in a large cohort of patients from the nationwide Veterans Affairs (VA) healthcare system. Our report, besides suggesting that cancer patients are more vulnerable to Covid-19 infection,^11,12^ also highlights the prevalence of Covid-19 and outcome of the disease based on racial characteristics, comorbidities, type of cancer and related treatment in a cohort of 22914 cancer patients.

## Methods

### Patients

This analysis was conducted using data from the VA Corporate Data Warehouse (CDW), which centralizes EHR data for patients seen at VA facilities nationwide. The study population is defined as veterans with cancer who were tested for Covid-19 at the VA. Cancer patients were identified as patients with at least one occurrence of an International Classification of Diseases (ICD) code for cancer between January 1, 2010 and May 4, 2020.^26^ Non-melanoma skin cancer and benign tumors were excluded. This study was performed under a protocol approved by the VA Boston Healthcare System Research and Development Committee.

### Study Variables and Outcomes

Patient demographics, Covid-19 laboratory tests, comorbidities, cancer type, cancer treatment, and outcomes were identified through structured data in the EHR. Patients were considered positive for Covid-19 if they had at least one positive test result, and negative if all Covid-19 test results were negative. Covid-19 tests that were cancelled or did not have a positive or negative result recorded were excluded from consideration. The Charlson comorbidity index and presence of individual comorbidities were derived from ICD codes in the year prior to each patient’s first Covid-19 test using the *comorbidity* package in R.^13^ BMI was calculated from each patient’s most recently recorded weight and height prior to their first Covid-19 test. Smoking status was defined using health factors.^14^ The geographical region of the VA hospital each patient was tested in, was extracted from structured data, with regions defined using the mapping in Supplementary Methods in Supplement 1. Cancer type was determined based on ICD codes in the study period, and patients may have multiple cancer types. Only cancer types with at least 200 patients are shown. The systemic therapies for cancer considered in this study were based on a list of approved cancer drugs tabulated by the NCI.^15^ Patients were identified as being on active or recent treatment for cancer at the time of Covid-19 testing if they received systemic therapy for cancer in the 6 months prior to their first test for Covid-19, and only treatments in this time frame were considered to determine the type of therapy received. As regards to outcomes, hospitalization was defined as any inpatient visit after the patient’s first Covid-19 test, and ICU admission was defined as a subset of hospitalizations where the specialty ward is either “Surgical ICU” or “Medical ICU”. Respiratory support was determined based on the presence of a current procedural terminology (CPT) or ICD procedure code for intubation (CPT 94002, 94003, 94004, 94005, ICD-10 Z99.1, Z99.11, Z99.12) or mechanical ventilation (CPT 31500, ICD-10 J95.851) after the patient’s first Covid-19 test. Missing data existed for race, ethnicity, and smoking status and was coded as a separate level in all analyses.

### Statistical Analysis

We evaluated the proportion of Covid-19 positive patients among those tested, along with 95% confidence intervals, and stratified by demographics, comorbidities, cancer treatment and cancer type. We also evaluated the proportion of patients experiencing each outcome among Covid-19 positive and negative patients, and defined the Covid-19 attributable risk of each outcome as the difference of these two proportions. We used chi-squared tests to assess differences in proportion. Odds ratio (OR) as to Covid-19 positive status by cancer type was assessed using univariate and multivariate logistic regression, adjusting for race, ethnicity, and smoking status in the multivariate models.

## Results

### Susceptibility to Covid-19 infection in cancer patients

We identified 22914 patients with history of cancer who were tested for Covid-19 infection on or before May 4, 2020, of whom 1794 patients (7.8%) reported positive **(Figure 1A)**. The prevalence of Covid-19 among those tested was similar across all ages (< 50 years to ≥80years; P =.158), although older patients with cancer were tested for Covid-19 more frequently **(eTable 1 in Supplement 1)**. Importantly, there was a significant difference in prevalence of infection across race and ethnicity, with 14.95% of African Americans (AA) but only 5.49% of white patients testing positive for Covid-19 (P<.001), and 10.87% of Hispanic/Latino patients vs 7.71% of non-Hispanic/Latino patients testing positive (P<.001). We also observed a significantly lower prevalence in cancer patients who were current smokers compared to former smokers or those who never smoked (5.26% vs 9.49%; P<.001). Compared to the overall sample (7.8% positive), there was a small but statistically significant negative association between susceptibility to Covid-19 and concomitant congestive heart failure (6.68% positive; P<0.001), peripheral vascular disease (6.51%; P<0.001), chronic obstructive pulmonary disease (6.27%; P<.001), and moderate/severe liver disease (5.15%; P =.023). Higher body mass index (BMI) was associated with increased prevalence of infection (P<.001).During the period of observation, Covid-19 was much more frequent in the North Atlantic (14.77%) than other regions (6.25% Continental, 7.85% Midwest, 2.37% Pacific, 3.21% Southeast; P<.001). Differences in susceptibility across race persisted within each region **(eTable 6)**.

**Figure 1:**
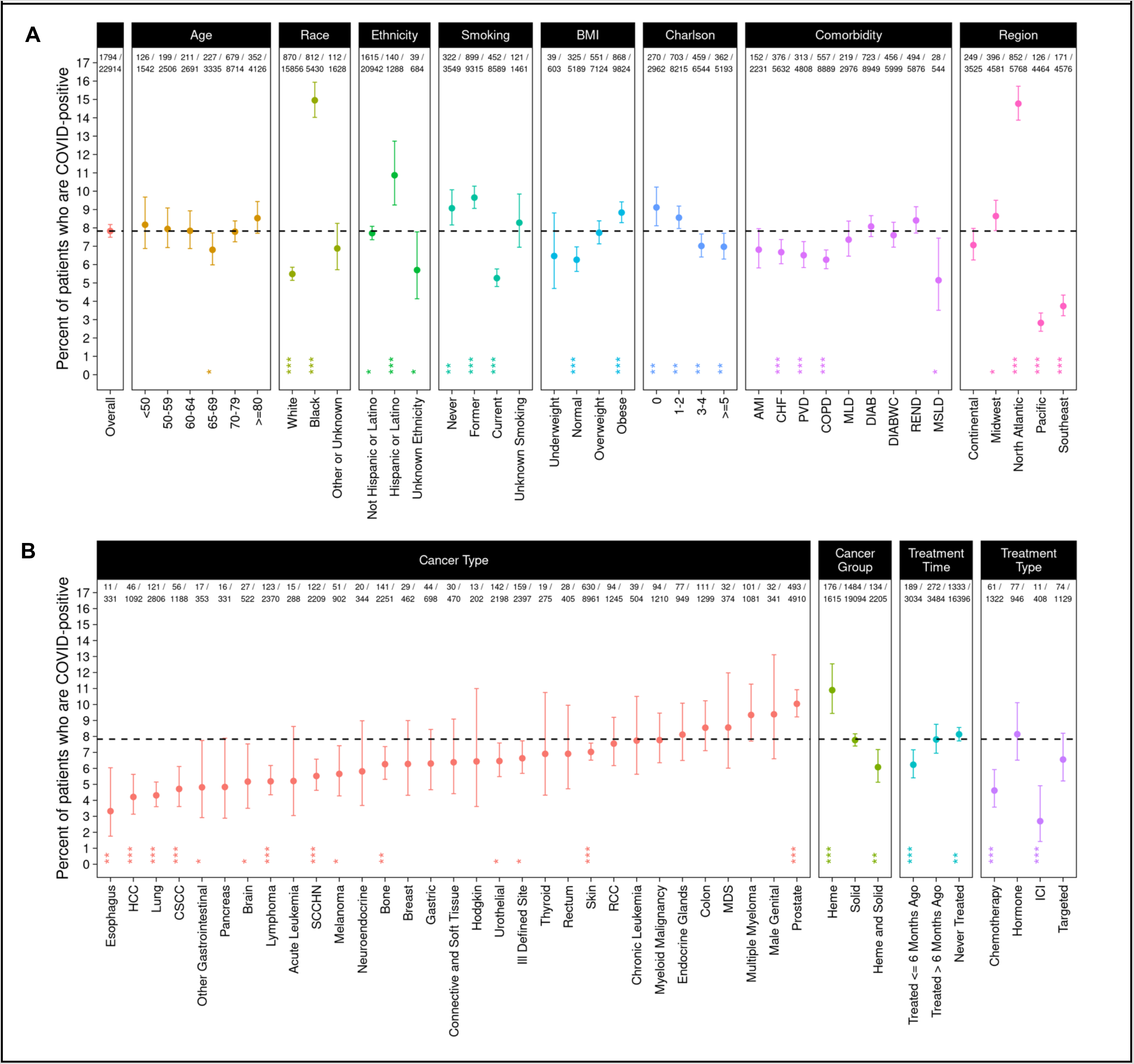
Covid-19 prevalence among cancer patients. Percent of Covid-19 positive patients among cancer patients tested for Covid-19, stratified by demographics, comorbidities, cancer type, and cancer therapy are represented. The dashed line indicates the overall percent positive (7.8%). The two rows at top show the number of Covid-19 positive patients, and the total number of cancer patients tested. In addition to the percent positive in each group, a 95% confidence interval and P value are shown. (* = P <.05; ** = P <.01; *** = P <.001).

We further investigated cancer types and prevalence of Covid-19 (**Figure 1B**) and observed higher prevalence in patients with hematologic malignancy (10.9%) in comparison to patients with solid tumors (7.8%, P<.001). Among specific cancer type, we observed significantly higher frequency of Covid-19 among patients with prostate cancer (10.0%, P< 0.001), while significantly lower frequency among patients with esophagus (3.3%, P =.003), hepatocellular carcinoma (HCC) (4.2%, P<.001), lung (4.3%, P<.001), cutaneous squamous cell carcinoma (CSSC) (4.7%, P<.001), lymphoma (5.2%, P<.001), squamous cell head and neck cancer (SCCHN) (5.5%, P<.001), bone cancer (6.3%, P =.004), and urothelial cancer (6.46%, P =.013), among others. To understand the relationship of race and cancer type, we assessed the odds of Covid-19 infection by cancer type, adjusting for race, ethnicity and smoking status **(eTable 2)**. Although the univariate analysis showed that patients with prostate cancer had a higher risk of Covid-19 infection, this was no longer the case after adjustment, suggesting the higher prevalence observed in prostate cancer may be affected by race, ethnicity, and/or smoking status. On the other hand, patients with esophageal cancer, HCC, lung cancer, lymphoma, SCCHN, bone cancer, melanoma, and other cancers continued to have a lower risk of Covid-19 infection even after adjusting for these factors.

Next, we evaluated the impact of active cancer treatment on susceptibility to Covid-19 infection and observed that patients receiving cancer therapy within the last 6 months had significantly lower prevalence (6.23%) of Covid-19 compared to those who received therapy > 6 months ago (7.81%) or never received therapy (8.13%; P =.002). Moreover, compared to the overall cohort (7.8%), patients receiving immune checkpoint inhibitors (2.70%, P<.001) or chemotherapy (4.61%, P<.001) within the last 6 months had significantly lower prevalence of Covid-19 while patients on hormonal therapy (8.14%, P =.763) or targeted therapy (6.55%, P =.114) did not have significantly different prevalence.

### Outcome of Covid-19 infection in cancer patients

We next evaluated the impact of Covid-19 infection on outcome in cancer patients and calculated the difference between frequency in positive vs negative as the Covid-19 attributable contribution. Overall, compared to Covid-19 negative cancer patients, Covid-19 positive cancer patients have higher frequency of hospitalizations (31.5% versus 43.8% respectively; an excess of 12.3% Covid-19 attributable), ICU admissions (7.8% vs 19.7% respectively; 11.9% Covid-19 attributable), respiratory support (1.3% vs 7.9% respectively; 6.6% Covid-19 attributable). Covid-19 attributable death was 10.9% with 14% death in Covid-19 positive compared to 3.1% in Covid-19 negative patients **(Figure 2)**.

**Figure 2:**
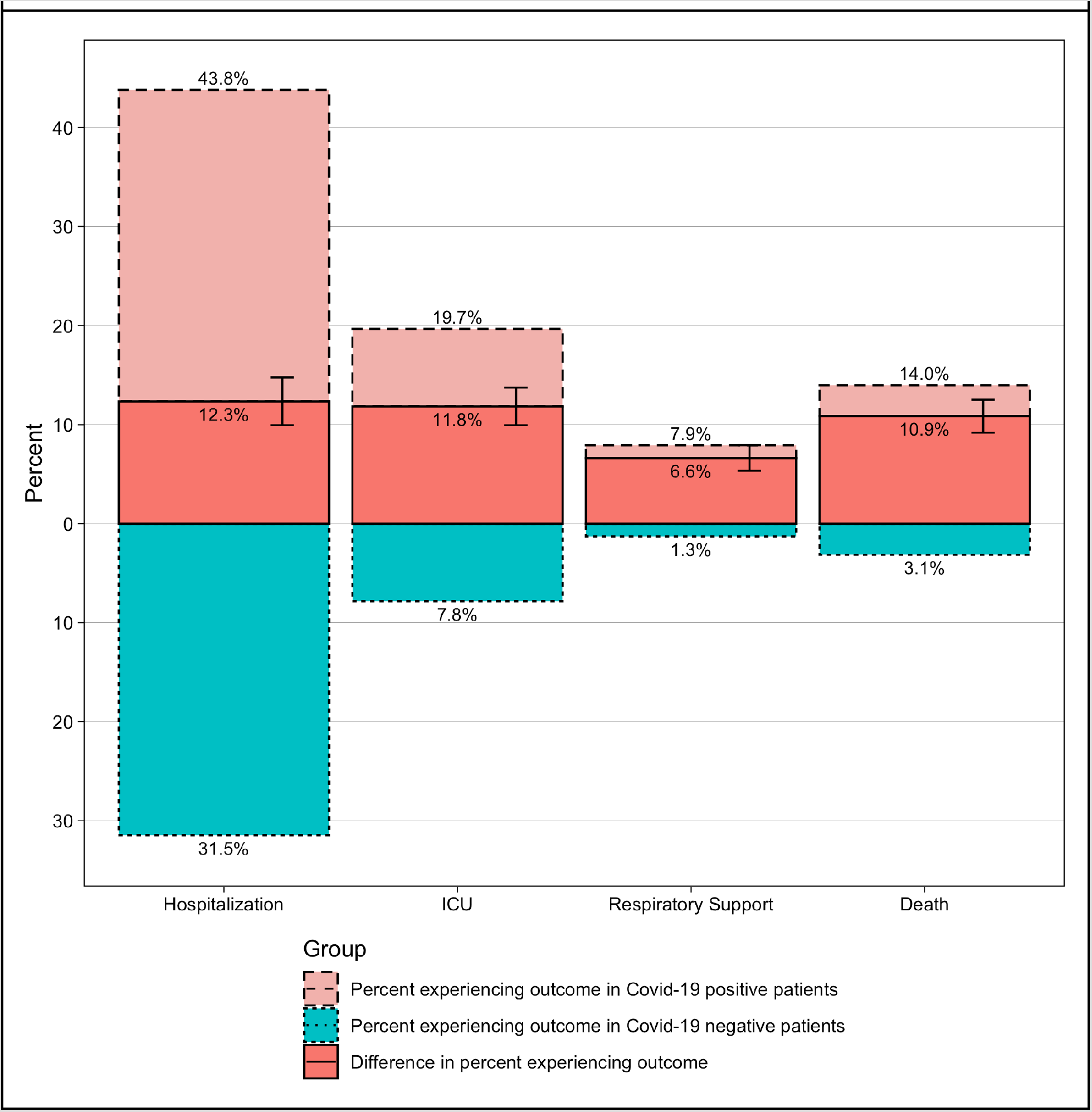
Outcome of cancer patients with Covid-19. Percent of patients experiencing hospitalization, ICU visits, respiratory support, and death in Covid-19 positive (blue) and negative (red) cancer patients are represented. The Covid-19 attributable risk of experiencing each outcome, i.e., the difference of the percent experiencing each outcome in Covid-19 positive compared to negative patients is also shown (darker red), along with 95% confidence intervals.

To further assess the impact of patient and disease-related features on Covid-19 attributable outcomes, we evaluated each of the demographic and comorbidity strata described above **(Figure 3A** and **Figure 4A)**. In general outcomes occur more frequently among Covid-19 positive cancer patients, however the difference attributable to Covid-19 infection varies widely across strata. Covid-19 attributable mortality is strongly associated with age, ranging from 0.23% among patients < 50 years old to 20.51% among patients ≥80 years old (P<.001). Presence of other comorbidities is also associated with increased Covid-19 attributable death, ranging from 3.07% among patients with Charlson score 0, to 14.96% among patients with Charlson score ≥5 (P<.001). Covid-19 attributable ICU admissions were more common in obese patients (14.34%) than in patients with normal BMI (5.87%, P<.001), as is need for respiratory support (8.94% vs 4.78%, P =.014); however mortality was not higher (10.45% in obese, 13.47% in normal; P =.21). Interestingly, Covid-19 attributable mortality was lower in current smokers (6.45%) compared to former/never smokers (11.99%; P =.002).

**Figure 3:**
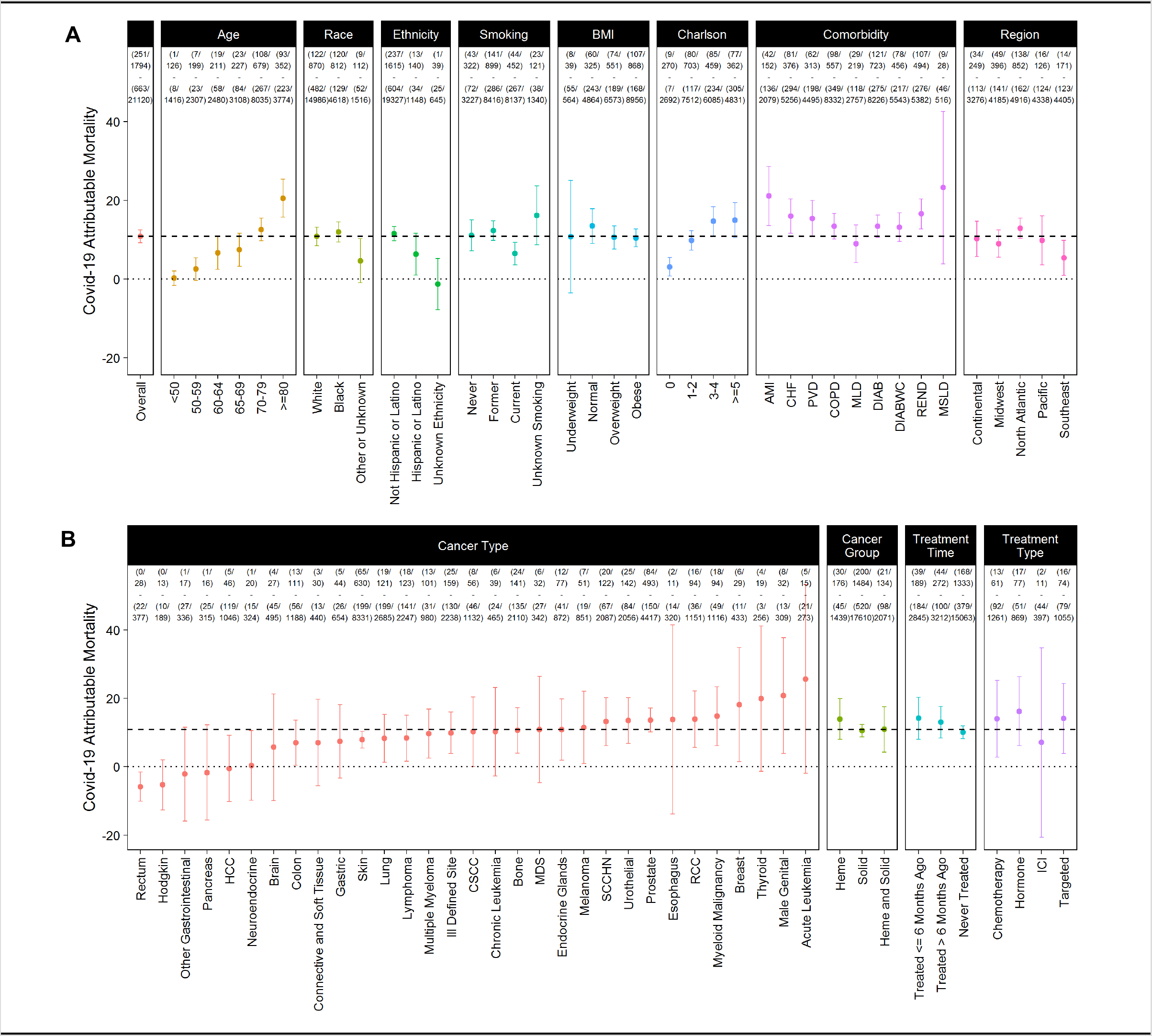
Covid-19 attributable mortality among cancer patients. Covid-19 attributable mortality defined as the difference in the percent mortality in Covid-19 positive compared to negative patients, is shown, along with 95% confidence intervals. The dashed line shows the Covid-19 attributable mortality in the overall cohort (14.4%), and the dotted line marks 0, the point where there is no Covid-19 attributable mortality. The four rows at top show the number of Covid-19 positive patients who died, the total number of Covid-19 positive patients, the number of Covid-19 negative patients who died, and the total number of Covid-19 negative patients.

**Figure 4:**
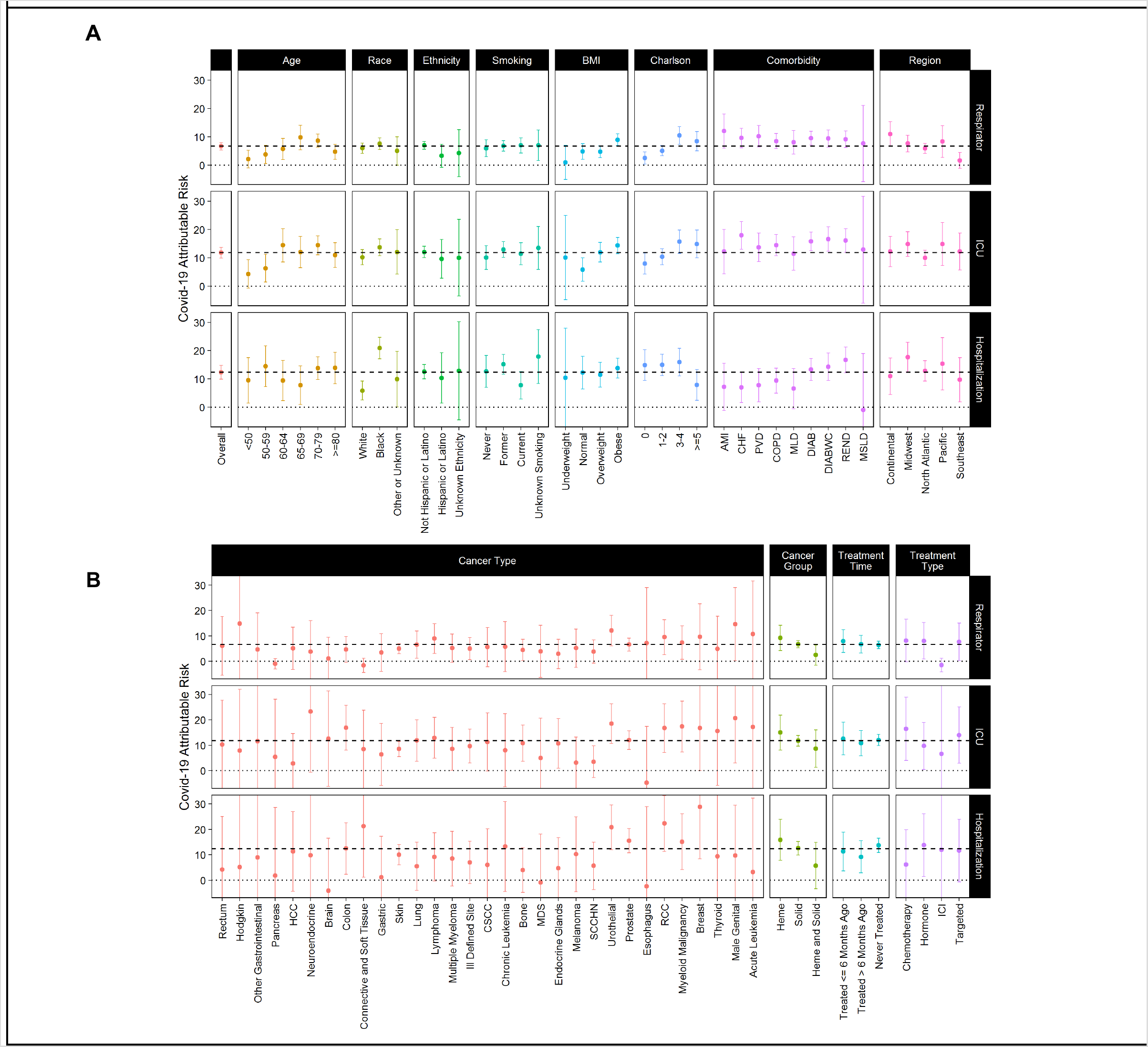
Covid-19 attributable hospitalizations, ICU admissions, and respiratory support among cancer patients. Covid-19 attributable hospitalizations, ICU admissions, and respiratory support defined as the difference in the percent of Covid-19 positive patients experiencing each outcome minus the percent of Covid-19 negative patients experiencing each outcome are shown. 95% confidence intervals are shown. The dashed line shows the Covid-19 attributable contribution in the overall cohort, and the dotted line marks 0, the point where there is no Covid-19 attributable contribution.

African Americans have increased Covid-19 attributable hospitalization (20.9%) compared to white patients (5.9%), a 3.5 fold difference (P<.001) **(Figure 4 and eTable 3)**. However, despite the higher Covid-19 prevalence as well as increased Covid-19 attributable hospitalization, we do not observe a significant difference in any of the other more serious Covid-19 attributable outcomes, including mortality (12.0% in AA vs 10.8% in white; P =.498), respiratory support (6.0% in AA vs 7.6% in white; P =.233), or ICU admissions (13.7% in AA vs 10.2% in white; P =.081). Similarly, we do not observe a significant difference in Covid-19 attributable outcomes for Hispanic/Latino compared to Non-Hispanic/Latino **(eTable 4)**.

Outcomes were variable based on type of cancer **(Figure 3B** and **Figure 4B)**. Covid-19 attributable deaths were observed in most but not all cancer types, with the highest attributable mortality in acute leukemia (25.64%), male genital (20.79%), and thyroid cancer (19.88%). Covid-19 attributable mortality is similar in patients recently treated for cancer (≤6 months ago, 14.17%) compared to those treated > 6 months ago (13.06%) or never treated with systemic therapy (10.09%; P =.250). There is a suggestive relationship between type of treatment and outcome attributable to Covid-19 infection. In patients receiving ICI within the last 6 months, Covid-19 attributable mortality (7.10%) is less than half that observed in patients receiving chemotherapy (14.02%), hormone therapy (16.21%), or targeted therapy (14.13%), though significance is not reached because of fewer observed deaths.

## Discussion

We investigated a large cohort of cancer patients evaluated for Covid-19 at the VA Healthcare System from across the United States in order to identify risk factors for Covid-19 susceptibility and outcomes in cancer patients. The overall frequency of Covid-19 positivity was 7.8% of tested patients. As of May 16th, 2020, according to the United States Center for Disease Control, the national positivity rate for Covid-19 infection in the US among patients aged of 18 and older is 13.5% with 126,627 positive patients out of 928,366 tested patients.^16^ Thus, the prevalence of Covid-19 among cancer patients appeared to be lower than in the general population. However, the real prevalence of Covid-19 remains uncertain, as a significant number of patients are not or have not been tested, particularly in the context of asymptomatic disease. Nevertheless, the observed lower prevalence of Covid-19 in cancer patients might be explained either by higher access to testing within the VA healthcare system and/or higher rates of testing in cancer patients in general.

Perhaps the most striking observation in our data is the significant racial and ethnic disparity in prevalence of Covid-19 infection. African-American (15%) and Hispanic (10.9%) cancer patients had significantly higher rates of Covid-19 infection in comparison to white cancer patients (5.5%), and also had higher rates of hospitalization. The biological and/or social basis for this observation, also recently reported in the general population,^17-19^ remains unclear. Overall, the observation of this disparity across all the regions of the country (**eTable 6**) rules out the possible role of regional conditions in explaining this difference. Covid-19 was more frequent among patients with prostate cancer which is also more frequent in African-American patients.^20^ Although after adjusting for race, ethnicity, and smoking status, this difference is no longer observed. The possible explanations requiring further investigation include socio-economic factors or genetic predisposition. Importantly, although AA cancer patients were more frequently admitted to the hospital, with equal access to care in the VA healthcare system, similar mortality rate was observed between AA and white cancer patients. Similar trend was also observed in Hispanic/Latino population. These results suggest distinct factors modulating Covid-19 susceptibility and the related-outcome.

The significantly reduced prevalence of Covid-19 infection in cancer patients with current smoking history, compared to those who have quit smoking or never smoked, is intriguing. This observation is also confirmed by observed reduced frequency of Covid-19 infection among patients with lung (4.31%, P< 0.001), SCCHN (5.52%, P< 0.001), and urothelial (6.46, P = 0.013) cancers, all malignancies associated with smoking. These differences might also be related to race or ethnicity **(eTables 3 and 4)**, or to socio-economics factors. An earlier preliminary report has also suggested association of active smoking with significantly lower rates of Covid-19 infections.^21^

This negative association requires further investigation. The role of nicotine, local epithelial cell changes or inflammatory environment may play a role.^22,23^ We did not observe significant impact of cardiopulmonary and other comorbidities as well as age of patients on rate of infection **(Figure 1)**. The lack of significant impact of the well-described influence of comorbidities on Covid-19 in our patients may suggest that pre-existence of cancer may outweigh the other comorbidities in regard to susceptibility to Covid-19. Moreover, it is possible that sick patients and patients under active cancer treatment may have been more cautious and thus less exposed to Covid-19, and this may affect the vulnerability. While we are unable to directly quantify severity of cancer illness in our dataset, we included two proxies for severity of disease that are available: (1) whether or not each patient was recently treated for cancer (≤6 months ago), treated > 6 months ago, or never treated with systemic therapy, which serves to some extent as a proxy for severity; and (2) the Charlson score which reflects severity of overall health condition prior to Covid-19 diagnosis. Our analysis does reveal significantly lower rate of infection in patients treated ≤6 months ago, though no significant difference in Covid-19 attributable mortality or other outcomes, and higher rates of infection in patients with higher Charlson score was observed.

Outcome of cancer patients infected by Covid-19 was characterized by higher rates of mortality. Although the frequency of Covid-19 positivity was lower in patients receiving cancer related therapy within 6 months of the Covid-19 infection, the overall mortality was not significantly different in comparison to no treatment or treatment prior to 6 months groups. These mortality rates are higher than the mortality rates reported in the global population^8,24^ and confirm higher vulnerability of cancer patients to Covid-19 infection. Therapies including conventional chemotherapy, targeted therapies with small molecules or monoclonal antibodies were associated with higher mortality rates in this study. Immune checkpoint inhibitor treatment was associated with a lower rate of infection however, only a small number of patients are in this group suggesting both caution in interpreting this data and also a need for further focused investigation in patients receiving ICI therapy and with Covid-19 positivity. The lower mortality rate in patients receiving this treatment confirms the recent report about the impact of this therapy on Covid-19 outcome.^25^ Although frequency of Covid-19 infection was not significantly different, higher rates of mortality were observed in elderly patients (19.5% in patients > 70 y.o.), patients with low Charlson score, underweight patients and patients with comorbidities, mainly cardio-vascular and renal disease **(Figure 3)**. While hematological malignancies were associated with higher rates of infection, similar Covid-19 attributable mortality was observed in comparison to solid malignancies. Fatality rate related to each cancer type was variable with breast, male genital, acute leukemia and thyroid cancer being associated with highest rate of Covid-19 related mortality **(Figure 3)**.

Our study has several limitations. First, the veteran population is primarily male and hence the study represents various trends and associations whose interpretations are restricted to male. Second, patients tested outside the VA system would not be included in this analysis. However, this number is likely to be very small as most patients who get cancer care in the VA do return for their healthcare needs to the VA. Third, it is possible that analyses are confounded by indication for testing. However, we believe that our comparison of those who tested positive to those who tested negative remains relevant, especially because Covid-19 testing criteria at VA hospitals was informed by centralized guidance from the VA Central Office, increasing consistency of testing criteria nationally.^27^

In conclusion, the presence of cancer changes the susceptibility to Covid-19 infection and affects overall outcome. The overall disease behavior is modulated by patient-related as well as cancer-related factors which needs to be considered in development of Covid-19 preventative strategies as well as modulation of cancer therapies to optimize the patient care. Importantly, having equal access to care is an important component to improving overall outcome.

## Data Availability

VA policy does not permit public sharing of the electronic health record data used in this study.

## Acknowledgements

This work was supported by the VA Office of Research and Development, Cooperative Studies Program (NRF, NVD, MTB), the VA Merit Review Award 1I01BX001584 (NCM), and NIH grants P01-155258-07 and P50-100707 (NCM). The views expressed are those of the authors and do not necessarily reflect the position or policy of the Department of Veterans Affairs or the United States government.

## Supplement 1 Supplementary Methods

### Geographical Region

The geographical region of the VA hospital each patient was tested in was extracted from structured data, with regions defined using the following mapping from the Corporate Data Warehouse:

- Continental (Arkansas, Colorado, Louisiana, Mississippi, Montana, Oklahoma, Texas, Utah, Wyoming)
- Midwest (Illinois, Indiana, Michigan, Minnesota, Missouri, Nebraska, North Dakota, Ohio, South Dakota, Wisconsin)
- North Atlantic (Connecticut, Delaware, District Of Columbia, Maine, Maryland, Massachusetts, New Hampshire, New Jersey, New York, North Carolina, Pennsylvania, Rhode Island, Vermont, Virginia, West Virginia)
- Pacific (Alaska, Arizona, California, Hawaii, Idaho, Nevada, New Mexico, Oregon, Washington)
- Southeast (Alabama, Florida, Georgia, Kentucky, Puerto Rico, South Carolina, Tennessee)

### Supplementary Results

**eTable 1.**
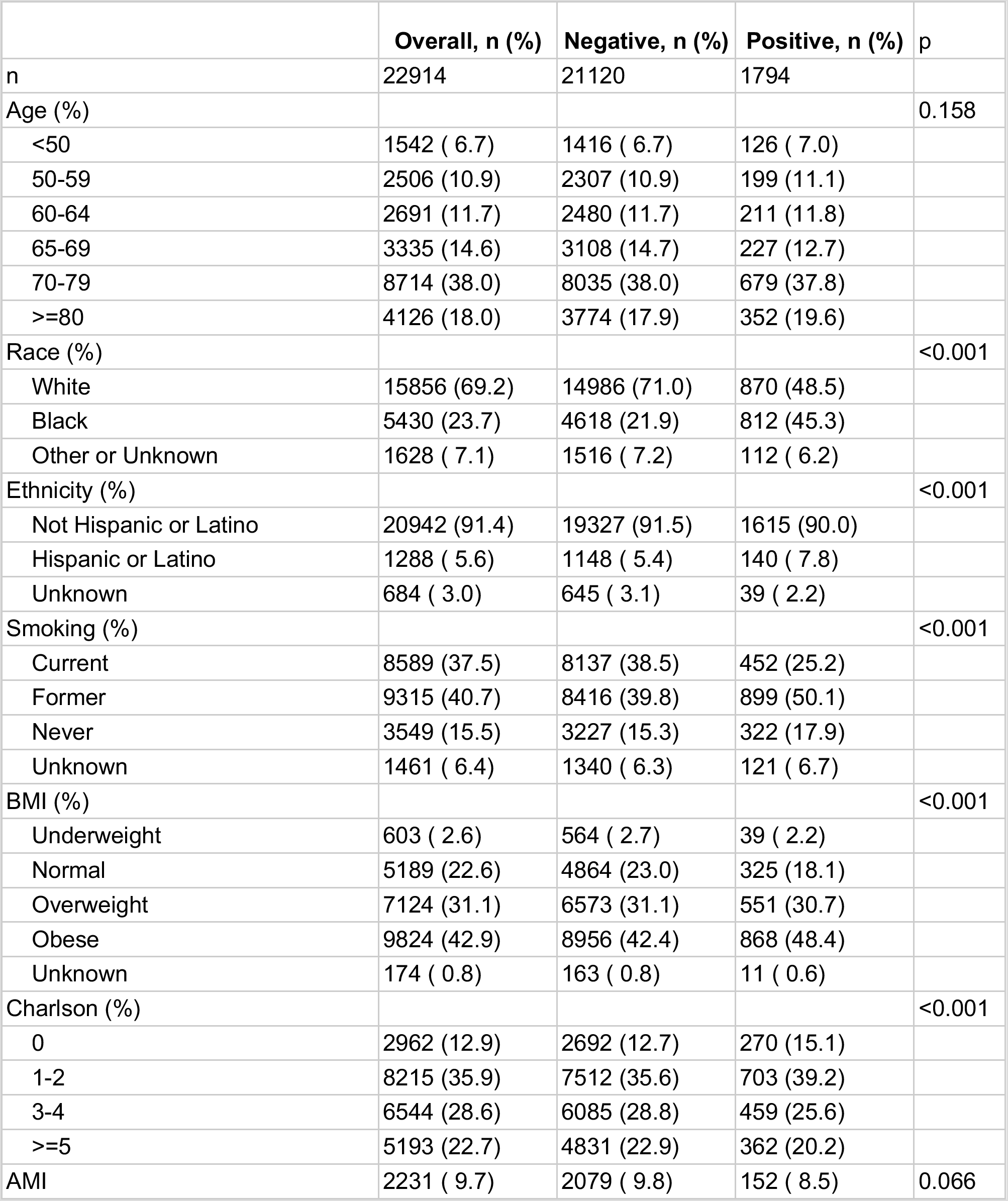
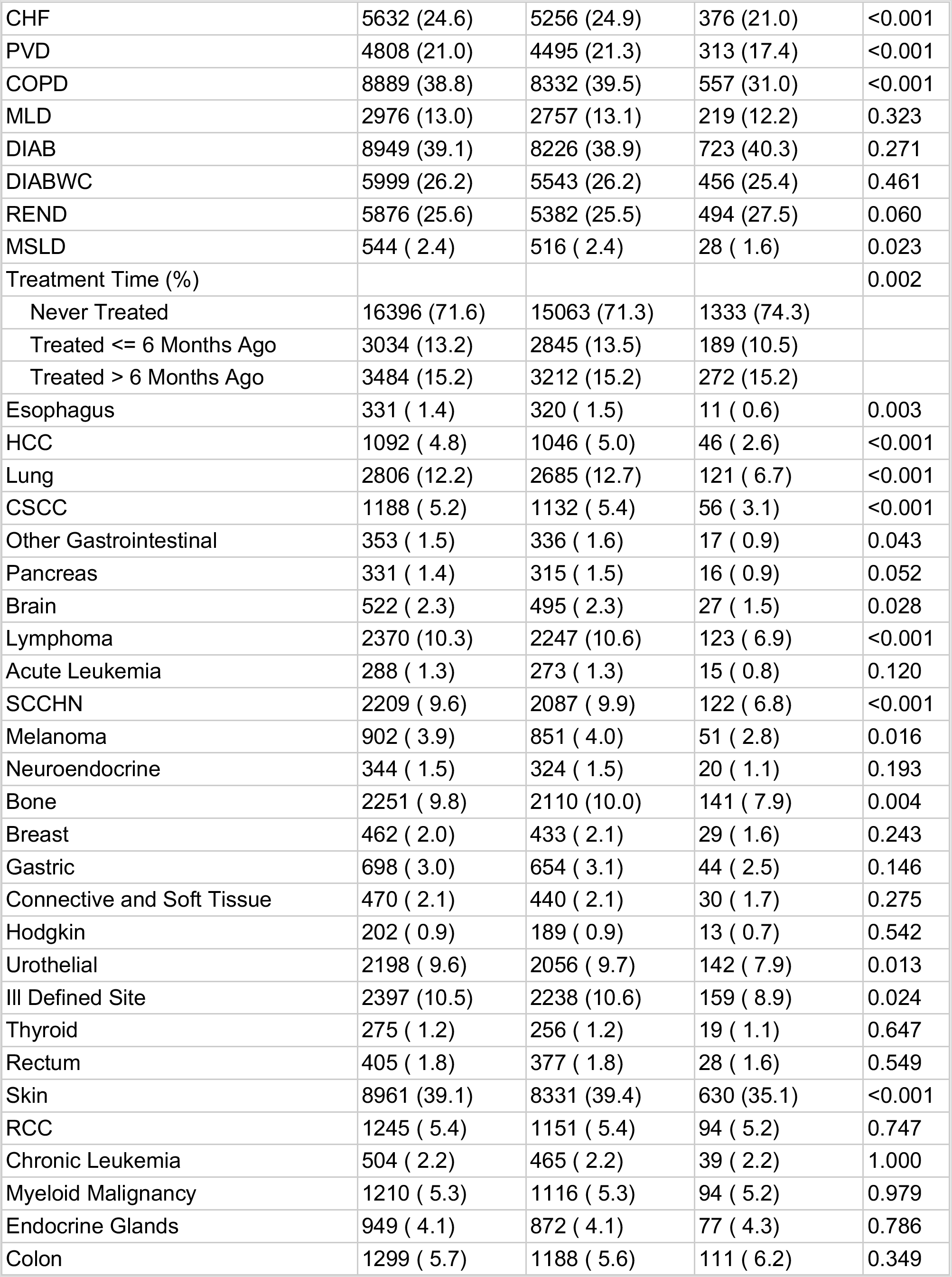
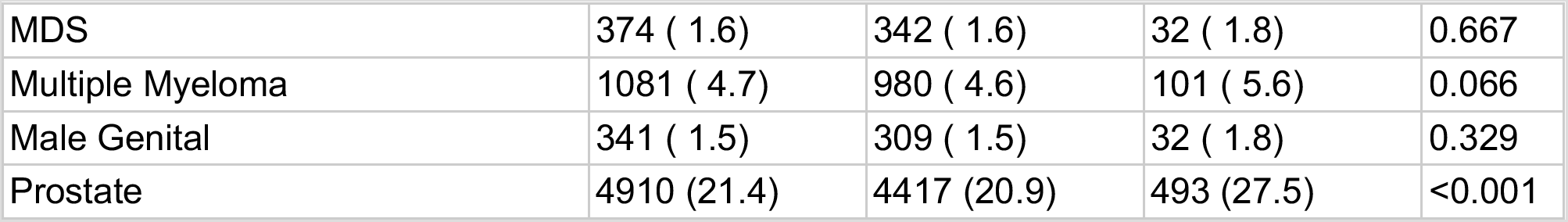
Patient characteristics in the full cohort of cancer patients tested for Covid-19, and stratified by patients who were negative or positive for Covid-19.

|  | Overall, n (%) | Negative, n (%) | Positive, n (%) | p |
| --- | --- | --- | --- | --- |
| n | 22914 | 21120 | 1794 |  |
| Age (%) |  |  |  | 0.158 |
| <50 | 1542 ( 6.7) | 1416 ( 6.7) | 126 ( 7.0) |  |
| 50-59 | 2506 (10.9) | 2307 (10.9) | 199 (11.1) |  |
| 60-64 | 2691 (11.7) | 2480 (11.7) | 211 (11.8) |  |
| 65-69 | 3335 (14.6) | 3108 (14.7) | 227 (12.7) |  |
| 70-79 | 8714 (38.0) | 8035 (38.0) | 679 (37.8) |  |
| >=80 | 4126 (18.0) | 3774 (17.9) | 352 (19.6) |  |
| Race (%) |  |  |  | <0.001 |
| White | 15856 (69.2) | 14986 (71.0) | 870 (48.5) |  |
| Black | 5430 (23.7) | 4618 (21.9) | 812 (45.3) |  |
| Other or Unknown | 1628 ( 7.1) | 1516 ( 7.2) | 112 ( 6.2) |  |
| Ethnicity (%) |  |  |  | <0.001 |
| Not Hispanic or Latino | 20942 (91.4) | 19327 (91.5) | 1615 (90.0) |  |
| Hispanic or Latino | 1288 ( 5.6) | 1148 ( 5.4) | 140 ( 7.8) |  |
| Unknown | 684 ( 3.0) | 645 ( 3.1) | 39 ( 2.2) |  |
| Smoking (%) |  |  |  | <0.001 |
| Current | 8589 (37.5) | 8137 (38.5) | 452 (25.2) |  |
| Former | 9315 (40.7) | 8416 (39.8) | 899 (50.1) |  |
| Never | 3549 (15.5) | 3227 (15.3) | 322 (17.9) |  |
| Unknown | 1461 ( 6.4) | 1340 ( 6.3) | 121 ( 6.7) |  |
| BMI (%) |  |  |  | <0.001 |
| Underweight | 603 ( 2.6) | 564 ( 2.7) | 39 ( 2.2) |  |
| Normal | 5189 (22.6) | 4864 (23.0) | 325 (18.1) |  |
| Overweight | 7124 (31.1) | 6573 (31.1) | 551 (30.7) |  |
| Obese | 9824 (42.9) | 8956 (42.4) | 868 (48.4) |  |
| Unknown | 174 ( 0.8) | 163 ( 0.8) | 11 ( 0.6) |  |
| Charlson (%) |  |  |  | <0.001 |
| 0 | 2962 (12.9) | 2692 (12.7) | 270 (15.1) |  |
| 1-2 | 8215 (35.9) | 7512 (35.6) | 703 (39.2) |  |
| 3-4 | 6544 (28.6) | 6085 (28.8) | 459 (25.6) |  |
| >=5 | 5193 (22.7) | 4831 (22.9) | 362 (20.2) |  |
| AMI | 2231 ( 9.7) | 2079 ( 9.8) | 152 ( 8.5) | 0.066 |
| CHF | 5632 (24.6) | 5256 (24.9) | 376 (21.0) | <0.001 |
| PVD | 4808 (21.0) | 4495 (21.3) | 313 (17.4) | <0.001 |
| COPD | 8889 (38.8) | 8332 (39.5) | 557 (31.0) | <0.001 |
| MLD | 2976 (13.0) | 2757 (13.1) | 219 (12.2) | 0.323 |
| DIAB | 8949 (39.1) | 8226 (38.9) | 723 (40.3) | 0.271 |
| DIABWC | 5999 (26.2) | 5543 (26.2) | 456 (25.4) | 0.461 |
| REND | 5876 (25.6) | 5382 (25.5) | 494 (27.5) | 0.060 |
| MSLD | 544 ( 2.4) | 516 ( 2.4) | 28 ( 1.6) | 0.023 |
| Treatment Time (%) |  |  |  | 0.002 |
| Never Treated | 16396 (71.6) | 15063 (71.3) | 1333 (74.3) |  |
| Treated <= 6 Months Ago | 3034 (13.2) | 2845 (13.5) | 189 (10.5) |  |
| Treated > 6 Months Ago | 3484 (15.2) | 3212 (15.2) | 272 (15.2) |  |
| Esophagus | 331 ( 1.4) | 320 ( 1.5) | 11 ( 0.6) | 0.003 |
| HCC | 1092 ( 4.8) | 1046 ( 5.0) | 46 ( 2.6) | <0.001 |
| Lung | 2806 (12.2) | 2685 (12.7) | 121 ( 6.7) | <0.001 |
| CSCC | 1188 ( 5.2) | 1132 ( 5.4) | 56 ( 3.1) | <0.001 |
| Other Gastrointestinal | 353 ( 1.5) | 336 ( 1.6) | 17 ( 0.9) | 0.043 |
| Pancreas | 331 ( 1.4) | 315 ( 1.5) | 16 ( 0.9) | 0.052 |
| Brain | 522 ( 2.3) | 495 ( 2.3) | 27 ( 1.5) | 0.028 |
| Lymphoma | 2370 (10.3) | 2247 (10.6) | 123 ( 6.9) | <0.001 |
| Acute Leukemia | 288 ( 1.3) | 273 ( 1.3) | 15 ( 0.8) | 0.120 |
| SCCHN | 2209 ( 9.6) | 2087 ( 9.9) | 122 ( 6.8) | <0.001 |
| Melanoma | 902 ( 3.9) | 851 ( 4.0) | 51 ( 2.8) | 0.016 |
| Neuroendocrine | 344 ( 1.5) | 324 ( 1.5) | 20 ( 1.1) | 0.193 |
| Bone | 2251 ( 9.8) | 2110 (10.0) | 141 ( 7.9) | 0.004 |
| Breast | 462 ( 2.0) | 433 ( 2.1) | 29 ( 1.6) | 0.243 |
| Gastric | 698 ( 3.0) | 654 ( 3.1) | 44 ( 2.5) | 0.146 |
| Connective and Soft Tissue | 470 ( 2.1) | 440 ( 2.1) | 30 ( 1.7) | 0.275 |
| Hodgkin | 202 ( 0.9) | 189 ( 0.9) | 13 ( 0.7) | 0.542 |
| Urothelial | 2198 ( 9.6) | 2056 ( 9.7) | 142 ( 7.9) | 0.013 |
| Ill Defined Site | 2397 (10.5) | 2238 (10.6) | 159 ( 8.9) | 0.024 |
| Thyroid | 275 ( 1.2) | 256 ( 1.2) | 19 ( 1.1) | 0.647 |
| Rectum | 405 ( 1.8) | 377 ( 1.8) | 28 ( 1.6) | 0.549 |
| Skin | 8961 (39.1) | 8331 (39.4) | 630 (35.1) | <0.001 |
| RCC | 1245 ( 5.4) | 1151 ( 5.4) | 94 ( 5.2) | 0.747 |
| Chronic Leukemia | 504 ( 2.2) | 465 ( 2.2) | 39 ( 2.2) | 1.000 |
| Myeloid Malignancy | 1210 ( 5.3) | 1116 ( 5.3) | 94 ( 5.2) | 0.979 |
| Endocrine Glands | 949 ( 4.1) | 872 ( 4.1) | 77 ( 4.3) | 0.786 |
| Colon | 1299 ( 5.7) | 1188 ( 5.6) | 111 ( 6.2) | 0.349 |
| MDS | 374 ( 1.6) | 342 ( 1.6) | 32 ( 1.8) | 0.667 |
| Multiple Myeloma | 1081 ( 4.7) | 980 ( 4.6) | 101 ( 5.6) | 0.066 |
| Male Genital | 341 ( 1.5) | 309 ( 1.5) | 32 ( 1.8) | 0.329 |
| Prostate | 4910 (21.4) | 4417 (20.9) | 493 (27.5) | <0.001 |

**eTable 2.**
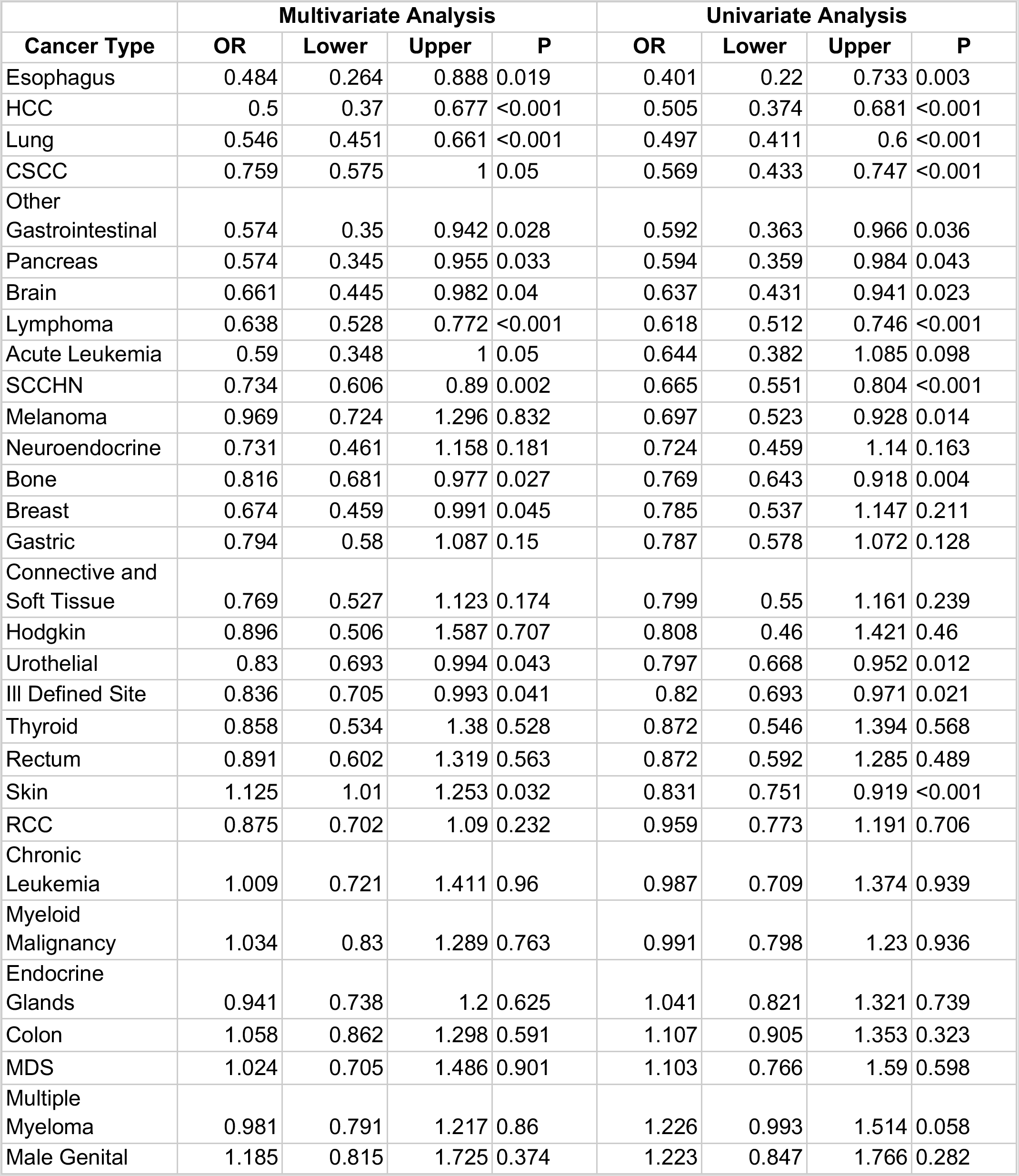
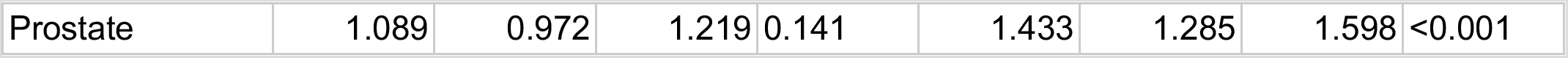
Odds ratio (OR) as to Covid-19 positive status by cancer type, in univariate logistic regression analysis and multivariate logistic regression, adjusting for race, ethnicity and smoking status. In addition to the odds ratio, the 95% confidence interval (lower, upper) and p value is shown.

**eTable 3.** Outcomes by Race and Covid-19 Result

|  | Outcome in Covid-19<br>Negative patients<br>n (%) |  | Outcome in Covid-19<br>Positive patients<br>n (%) |  | Covid-19<br>attributable<br>outcome % |  | Ratio of<br>Covid-19<br>attributable<br>outcome in<br>AA/White |
| --- | --- | --- | --- | --- | --- | --- | --- |
|  | White | AA | White | AA | White | AA |  |
| Hospitalization | 4835 (32.3) | 1365 (29.6) | 332 (38.2) | 410 (50.5) | 5.9 | 20.9 | 3.5 |
| ICU | 1204 (8.0) | 350 (7.6) | 159 (18.3) | 173 (21.3) | 10.3 | 13.7 | 1.3 |
| Respiratory Support | 191 (1.3) | 60 (1.3) | 63 (7.2) | 72 ( 8.9) | 5.9 | 7.6 | 1.3 |
| Death | 482 (3.2) | 129 (2.8) | 122 (14.0) | 120 (14.8) | 10.8 | 12 | 1.1 |

**eTable 4.** Outcomes by Ethnicity and Covid-19 Result

|  | Outcome in Covid-19<br>Negative patients<br>n (%) |  | Outcome in Covid-19<br>Positive patients<br>n (%) |  | Covid-19 attributable<br>outcome % |  | Ratio of Covid-19 attributable<br>outcome in<br>Hispanic or<br>Latino/Not<br>Hispanic or<br>Latino |
| --- | --- | --- | --- | --- | --- | --- | --- |
|  | Not Hispanic<br>or Latino | Hispanic<br>or Latino | Not<br>Hispanic<br>or Latino | Hispanic<br>or Latino | Not<br>Hispanic<br>or Latino | Hispanic<br>or Latino |  |
| Hospitalization | 6123 (31.7) | 324 (28.2) | 715 (44.3) | 54 (38.6) | 12.6 | 10.4 | 0.8 |
| ICU | 1518 ( 7.9) | 86 ( 7.5) | 322 (19.9) | 24 (17.1) | 12 | 9.6 | 0.8 |
| Respiratory<br>Support | 244 ( 1.3) | 20 ( 1.7) | 133 ( 8.2) | 7 ( 5.0) | 6.9 | 3.3 | 0.5 |
| Death | 604 ( 3.1) | 34 ( 3.0) | 237 (14.7) | 13 ( 9.3) | 11.6 | 6.3 | 0.5 |

**eTable 5.** Outcomes by Tumor Type and Covid-19 Result

|  | Outcome in Covid-19 Negative patients<br>n (%) |  |  | Outcome in Covid-19 Positive patients<br>n (%) |  |  | Covid-19 attributable outcome % |  |  |
| --- | --- | --- | --- | --- | --- | --- | --- | --- | --- |
|  | Solid | Heme | Solid and Heme | Solid | Heme | Solid and Heme | Solid | Heme | Solid and Heme |
|  | 17610 | 1439 | 2071 | 1484 | 176 | 134 |  |  |  |
| Hospitalization | 5211 (29.6) | 548 (38.1) | 886 (42.8) | 626 (42.2) | 95 (54.0) | 65 (48.5) | 12.6 | 15.9 | 5.7 |
| ICU | 1283 (7.3) | 135 (9.4) | 237 (11.4) | 283 (19.1) | 43 (24.4) | 27 (20.1) | 11.8 | 15 | 8.7 |
| Respiratory Support | 206 (1.2) | 23 (1.6) | 41 (2.0) | 117 ( 7.9) | 19 (10.8) | 6 (4.5) | 6.7 | 9.2 | 2.5 |
| Death | 520 (3.0) | 45 (3.1) | 98 (4.7) | 200 (13.5) | 30 (17.0) | 21 (15.7) | 10.5 | 13.9 | 11 |

**eTable 6.**
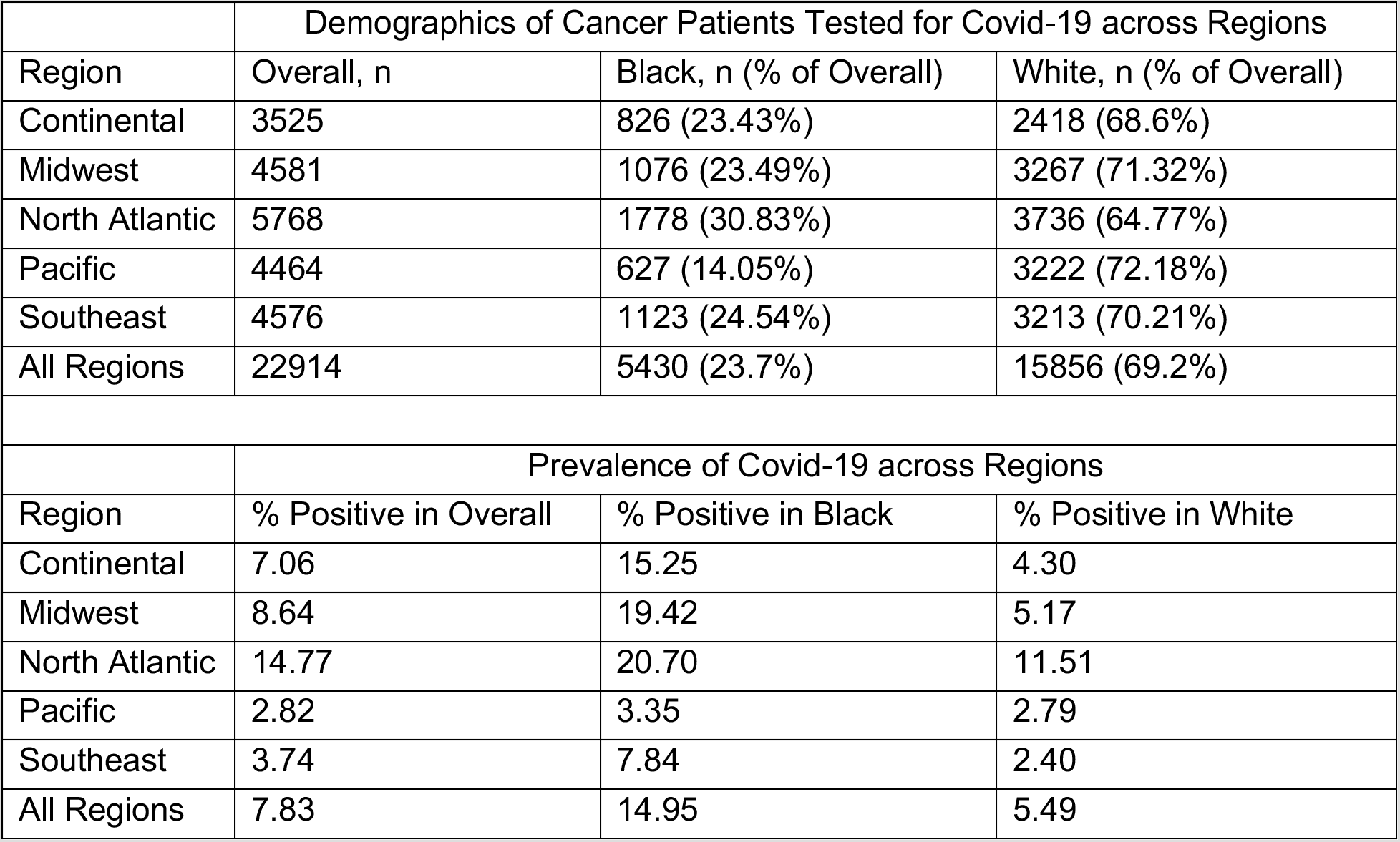
Regional data on Covid-19 prevalence and race. Higher prevalence is observed in black patients across all regions

